# Nonpharmaceutical Interventions Remain Essential to Reducing COVID-19 Burden Even in a Well-Vaccinated Society: A Modeling Study

**DOI:** 10.1101/2021.03.29.21254568

**Authors:** Tomás M. León, Jason Vargo, Erica S. Pan, Seema Jain, Priya B. Shete

## Abstract

Vaccination and non-pharmaceutical interventions (NPIs) reduce transmission of SARS-CoV-2 infection, but their effectiveness depends on coverage and adherence levels. We used scenario modeling to evaluate their effects on cases and deaths averted and herd immunity. NPIs and vaccines worked synergistically in different parts of the pandemic to reduce disease burden.

## Background

Since the beginning of the pandemic, SARS-CoV-2 infections have led to a reported 2,492,668 cases and 40,361 deaths through December 31, 2020 in California. Amidst the winter surge, a regional stay-at-home order was in effect at the end of 2020 that affected 98% of Californians. Concurrently, starting in December, administration of two authorized vaccines began in the state in accordance with recommended prioritization schemes, first targeting the Phase 1A priority groups [1-6]. In January, California then relaxed restrictions to allow adults 65+, who are at highest risk of COVID-related mortality, to receive the vaccine sooner [6]. The secondary prioritization based on age was due in part to data from studies of the two vaccines describing higher efficacy for reducing disease severity as measured by hospitalization and death [2-5].

The potential synergistic effects of vaccination and non-pharmaceutical interventions (NPIs) to reduce SARS-CoV-2 transmission and consequent disease are not well understood. Considering the complexity of SARS-CoV-2 transmission dynamics and health effects, it is important to estimate the effects of vaccine uptake alongside other interventions using mathematical modeling. We aimed to assess the effects of different levels and strategies of NPIs and vaccination on key epidemiological outcomes in California, specifically cases, deaths, and time until herd immunity. We investigated the impact on projected cases and deaths of a 65+ vaccine prioritization strategy compared with vaccinating all adults, and considered the interaction between vaccines that partially reduce transmission with continued NPIs on these outcomes for the population of California. Finally, we examined how these combinations of NPIs and vaccination levels might impact progression to a theoretical herd immunity in 2021.

## Methods

We used a modified, age-stratified compartmental (SEIR) model based on a previously published model [7] and parameterized for the state of California. We varied adherence to NPIs by scaling of the contact matrix between age groups, including 65 – 74 and 75 years and older to account for changing age-based prioritization of vaccines. The effects of specific NPIs were not separately estimated, but rather the rescaled contact matrix was used to represent the total effect of different NPIs and adherence levels on contact rates for specified scenarios. Initial conditions corresponded to PCR-confirmed cases of COVID-19 and consequent deaths reported to the California Department of Public Health by January 1, 2021. We assumed that cases diagnosed in the preceding 2 weeks were actively infectious and cases diagnosed in the following two weeks after January 1 were exposed as of January 1 but not yet infectious. Immunity from prior infection (i.e., those considered “Recovered”) at the start of model simulations was estimated from available statewide seroprevalence data, which suggested that a third of estimated infections became cases confirmed by PCR. Theoretical “herd immunity” threshold is considered 60 – 70% based on an estimated *R*_*0*_ of 2.5 – 3.3. To estimate herd immunity, we included all successful vaccinations and natural infections and did not consider waning immunity.

### Scenarios

We compared 18 different scenarios covering 3 NPI levels (implemented as contact rates), 3 vaccine coverage levels, and 2 vaccine prioritization strategies to model their interactive effects on COVID-19 cases, deaths, and herd immunity. Vaccination coverage rate varied between 20%, 40% and 60% of the total California population (40.3 million) over 6 months. Contact rates (i.e., the amount of contact between persons and across age strata that could facilitate COVID-19 transmission) varied by scaling the contact matrix for low (corresponding to the 2019 pre-pandemic rates), moderate (between June and October 2020 when mobility was fairly stable), and high (April 2020, after the first stay-at-home order) NPI scenarios [8]. Vaccination prioritization scenarios considered were either all adults (18+) or adults 65+ only, according to the implemented prioritization scheme in California.

We assumed that the currently approved vaccines work similarly well and are 90% effective at preventing severe disease, hospitalization, and death, and 50% effective at reducing transmission [9]. We assumed 25% vaccine hesitancy in every age stratum. Vaccine distribution occurred at a constant daily rate according to the level of coverage/speed specified in each scenario such that distribution was completed over the six months from January through June 2021. The vaccines are modeled as providing “all-or-nothing” protection and with variable transmission blocking as previously described [7].

## Results

Scenarios of different NPI levels with different levels of vaccine coverage avert more cases and deaths compared with a base scenario of pre-pandemic contact rates and no vaccination (Figure 1). For cases, NPI adherence had the biggest effect on reducing incidence compared with the vaccination-related factors, particularly in the first half of the simulation (January-April). For cases, scenario results ranged from 450,000 (9%) cases averted (65+ prioritized, 20% vaccine coverage, low NPIs) to 3,850,000 (79%) cases averted (all adults, 60% vaccine coverage, high NPIs) (Supplement Table 1). For deaths, vaccination-related factors and NPIs interacted synergistically to reduce deaths, with 65+ prioritization having a significant impact. For deaths, scenario results range from 21,300 (11%) deaths averted (all adults, 20% vaccine coverage, low NPIs) to 128,700 (68%) deaths averted (65+ prioritized, 60% vaccine coverage, high NPIs) (Supplemental Table 2).

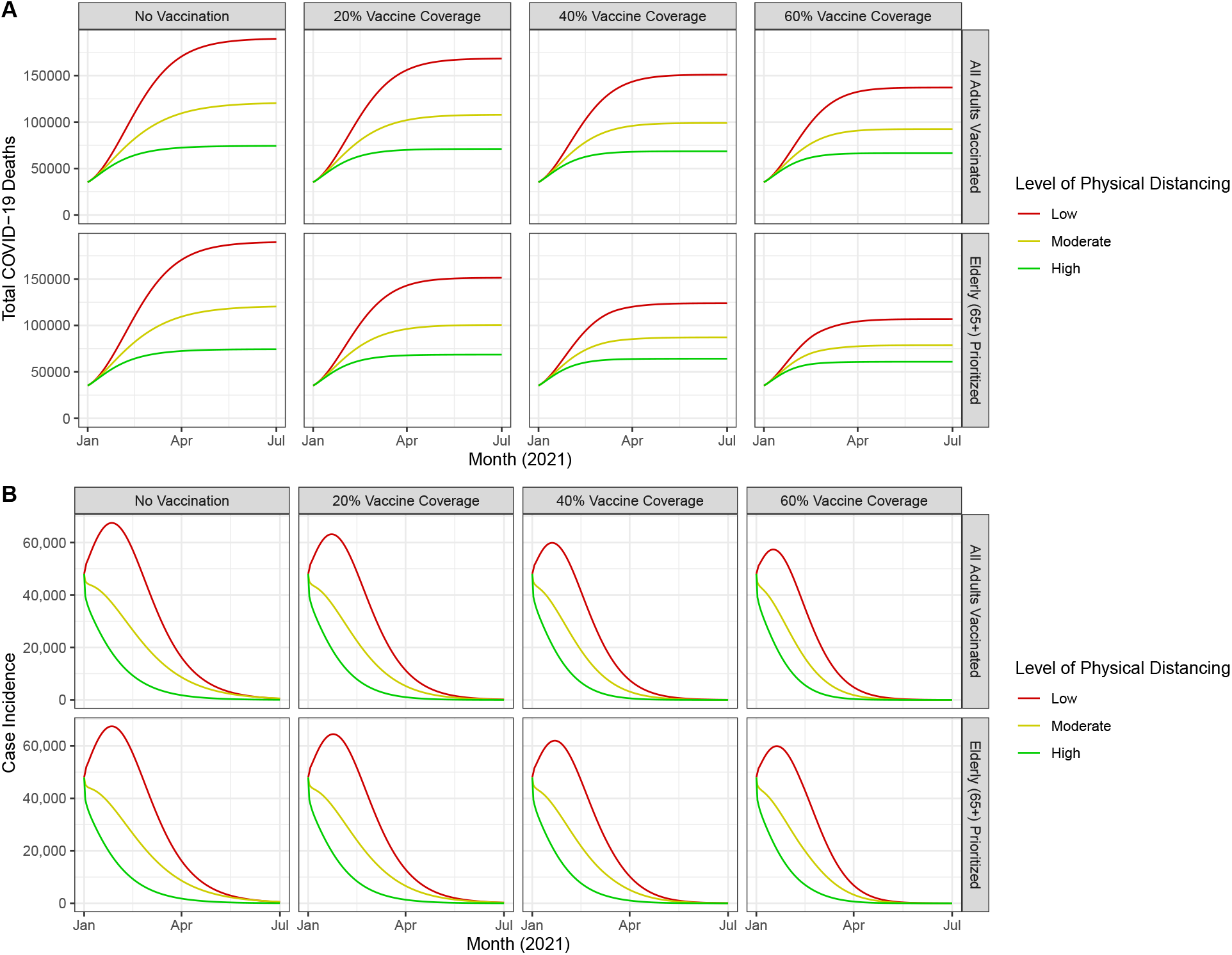

Model results for progression towards herd immunity over the course of the projection period vary by scenario (Figure 2). With 60% intended vaccination coverage, herd immunity is reached between May and July 2021 in California. Herd immunity is not reached in any of the 0% and 20% vaccination coverage scenarios. While high levels of NPI adherence delay achievement of herd immunity compared with lower levels of NPI adherence by 2 months at 60% vaccination coverage, this scenario averts 2,540,000 more cases (68%) and 45,800 more deaths (36%).

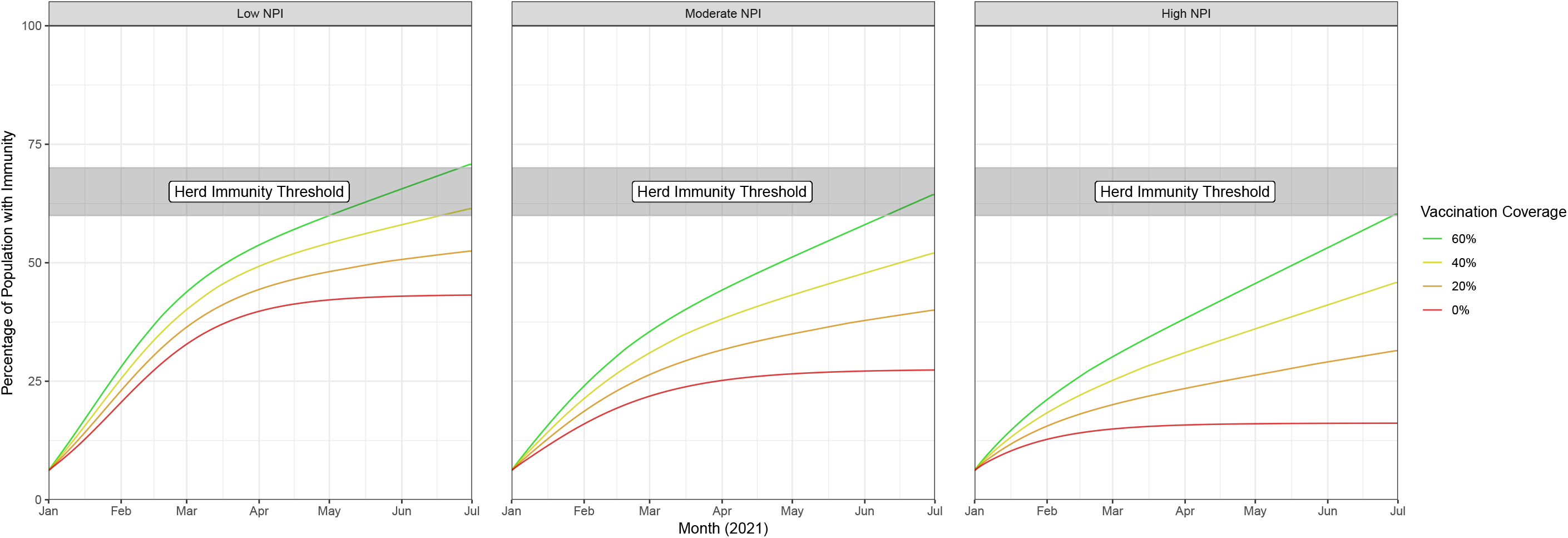

## Discussion

Our results show that even with high vaccination coverage, concomitant NPIs are required to reduce cumulative deaths (Figure 1A). NPI adherence figures prominently because NPIs drive total cases down faster than vaccinations alone (Figure 1B). Even with no vaccination, a high level of NPI adherence averts 74% of COVID-19 cases and 61% of COVID-19 deaths over the simulation period between January and July 2021 (Supplemental Table 1). Our findings demonstrate that continued NPIs are important for preventing additional deaths and cases while vaccine deployment scales up. In a scenario in which 20% of the total population has been vaccinated with age prioritization, a moderate level of physical distancing would still avert 89,000 (47%) deaths compared to low physical distancing, which would avert 38,300 (20%) deaths. Current COVID-19 case and death trajectories in 2021 suggest that California is operating at moderate NPIs scenario levels, and our current vaccination strategy prioritizes the +65 cohort. Currently, California is on track to vaccinate 60% or more of the population by July 2021.Under these conditions we would expect to avert 59% of deaths compared to the base scenario.

Our model also describes the relationship between vaccination coverage, NPIs and “herd immunity,” the level of immunity sufficient to disrupt and prevent sustained disease transmission. Even with accelerating vaccination rates, a level of herd immunity sufficient to eliminate the need for NPIs may not be achieved for 6 months or more. Although maintaining NPIs during vaccination scale-up may be challenging, such measures could prevent 100,000+ additional deaths in California from occurring. The effect of higher NPI adherence on delaying herd immunity, due to averted infections contributing natural immunity, is outweighed by the effects of increasing vaccination speed and coverage on reducing deaths and incident cases (Figure 2).

Our results suggest that prioritizing Californians 65+ reduces cumulative deaths substantially at all levels of vaccine coverage. Based on our findings, we estimate that 40% vaccine coverage in the 65+ age group has approximately the same reduction in deaths as vaccination coverage of at least 80% among the entire adult population over the same time. While not explicitly modeled, hospitalizations preceding deaths are also likely to be substantially reduced by prioritizing adults 65+, which would otherwise strain California’s health system.

Like all scenario models, ours has limitations, the first being inherent uncertainty about parameters and scenario conditions. Our assumptions about the effects of vaccination on transmission are conservative, due to relatively limited evidence of effectiveness against asymptomatic transmission. This may also affect the timeframe for reaching herd immunity. Additionally, our analyses do not currently account for increasing prevalence of novel SARS-CoV-2 variants that are more infectious or immune evasive. Each of these challenges support maintaining a conservative approach to NPIs.

Our analyses provide evidence to support continued disease control and prevention efforts, including masking, social/physical distancing, ventilation, and hand hygiene (i.e., NPIs), during initial COVID-19 vaccine implementation. First, the effect of vaccination in low NPI scenarios is attenuated by ongoing transmission while herd immunity is still distant. Second, despite potential accelerated scale up of vaccinations, levels of herd immunity required to reduce the need for NPIs would not be reached until summer 2021 or later, absent a major increase in vaccine supply and distribution.

Public health stakeholders should continue implementing both vaccines and physical distancing measures (NPIs) simultaneously to reduce transmission, hospitalizations, and deaths as we aspire for pandemic control and until herd immunity, driven by vaccination, reaches a sufficient level to ensure protection for the most vulnerable.

## Data Availability

California COVID-19 data used in this modeling study is publicly available through https://covid19.ca.gov/.

## Acknowledgments/Disclosures

The views and opinions expressed by the authors are their own and do not necessarily represent the views and opinions of the California Department of Public Health, the California Health and Human Services Agency, or the Administration. We would like to thank Kate Bubar and Daniel Larremore for helpful comments on this manuscript and Megha Mehrotra for support with seroprevalence estimates.

## Funding Statement

This work was supported by the California Department of Public Health.

There are no public meetings at which this information has been previously presented.

