## Supplemental Tables for "Nonpharmaceutical Interventions Remain Essential to Reducing COVID-19 Burden Even in a Well-Vaccinated Society: A Modeling Study"

**Supplemental Table 1:** Cumulative cases averted (counts and percentages) during the projection period. The base-case scenario assumes liberalizing of NPIs and high levels of contact and mixing between individuals in the absence of any vaccine coverage. In this scenario an estimated 4,900,000 cases would occur.

| **All Adults (18+) Vaccinated** | | | | |
| --- | --- | --- | --- | --- |
|  | **0% Vax Coverage** | **20% Vaccine Coverage** | **40% Vaccine Coverage** | **60% Vaccine Coverage** |
| **Low NPIs** | Base scenario | 680,000  (-13.9%) | 1,230,000  (-25.3%) | 1,670,000  (-34.4%) |
| **Moderate NPIs** | 2,090,000  (-42.9%) | 2,510,000  (-51.7%) | 2,810,000  (-57.8%) | 3,030,000  (-62.3%) |
| **High NPIs** | 3,570,000  (-73.5%) | 3,690,000  (-75.9%) | 3,780,000  (-77.7%) | 3,850,000  (-79.2%) |
| **Adults 65+ Prioritized for Vaccination** | | | | |
|  | **0% Vax Coverage** | **20% Vaccine Coverage** | **40% Vaccine Coverage** | **60% Vaccine Coverage** |
| **Low NPIs** | Base scenario | 450,000  (-9.3%) | 840,000  (-17.3%) | 1,220,000  (-25.1%) |
| **Moderate NPIs** | 2,090,000  (-42.9%) | 2,350,000  (-48.4%) | 2,570,000  (-52.9%) | 2,770,000  (-57.0%) |
| **High NPIs** | 3,570,000  (-73.5%) | 3,650,000  (-75.0%) | 3,710,000  (-76.2%) | 3,760,000  (-77.4%) |

**Supplemental Table 2:** Cumulative deaths averted (counts and percentages) during the projection period. The base-case scenario assumes liberalizing of NPIs and high levels of contact and mixing between individuals in the absence of any vaccine coverage. In this scenario an estimated 190,000 deaths would occur.

| **All Adults (18+) Vaccinated** | | | | |
| --- | --- | --- | --- | --- |
|  | **0% Vax Coverage** | **20% Vaccine Coverage** | **40% Vaccine Coverage** | **60% Vaccine Coverage** |
| **Low NPIs** | Base scenario | 21,300  (-11.2%) | 38,600  (-20.3%) | 52,500  (-27.7%) |
| **Moderate NPIs** | 69,300  (-36.5%) | 81,800  (-43.1%) | 90,600  (-47.8%) | 97,200  (-51.3%) |
| **High NPIs** | 115,300  (-60.8%) | 118,600  (-62.6%) | 121,100  (-63.9%) | 123,100  (-64.9%) |
| **Adults 65+ Prioritized for Vaccination** | | | | |
|  | **0% Vax Coverage** | **20% Vaccine Coverage** | **40% Vaccine Coverage** | **60% Vaccine Coverage** |
| **Low NPIs** | Base scenario | 38,300  (-20.2%) | 65,700  (-34.6%) | 82,900  (-43.7%) |
| **Moderate NPIs** | 69,300  (-36.5%) | 89,100  (-47.0%) | 102,400  (-54.0%) | 111,000  (-58.5%) |
| **High NPIs** | 115,300  (-60.8%) | 121,100  (-63.8%) | 125,500  (-66.2%) | 128,700  (-67.9%) |
